# Processed food intake assortativity in the personal networks of older adults

**DOI:** 10.1101/2024.01.25.24301787

**Authors:** Marian-Gabriel Hâncean, Jürgen Lerner, Matjaž Perc, José Luis Molina, Marius Geantă, Iulian Oană, Bianca-Elena Pin□Doiu-Mihăilă

## Abstract

Existing research indicates that dietary habits spread through social networks, yet the impact on older adults in Eastern Europe, particularly in rural areas, is less understood. We examine the influence of social networks on the consumption of high-salt processed foods among older adults in rural Romania. Using a personal network analysis design, we analyzed data from 83 participants and their social contacts with multi-level regression models. Our findings reveal assortativity in dietary habits among network ties, underlining the need for interventions that consider the broader social context to effectively mitigate high salt intake and its health risks in this population.

## 1. Introduction

Available evidence substantiates that eating patterns shape and, at the same time, are shaped by social networks [1]. Some argue that specific eating behaviors are the result of social influence. Namely, food habits circulate through social relationships, e.g., strong ties lead to similarities in food preferences [2], workplace networks impact employees’ dietary choices [1], and men’s relationships can influence dietary behavior and weight loss [3]. However, food habits are not merely the result of peer influence processes, norms, or social networks [4]. Cultural tastes and preferences (eating included) can also shape personal networks [5]. Thus, we can identify socialization (peer influence), homophily (social selection), and environmental confounding [6] as social mechanisms underpinning human behavior and, implicitly, dietary patterns.

Health behaviors and states can exhibit clustering within social networks [7]. Individuals’ health correlates with the health of their social connections, such as friends, siblings, spouses, and neighbors. Social networks may be crucial in understanding human practices such as food choices or health-related behaviors [8]. Nevertheless, network-oriented research work is still coalescing in this area, being in a nascent state. The few existent network studies are limited in their scope. Their focus is on adolescents [9–11] or population samples of friends and family [1, 12]. Therefore, generalizing findings to other age groups and cultural spaces is challenging.

Furthermore, in the case of older adults, there is mixed evidence concerning the association between social network type and dietary behavior [13]. Also, even if social relationships are generally linked to dietary behavior, it is unclear which structural aspects of personal networks affect healthful dietary behavior [12]. The investigation of how social networks influence eating habits is at a very early stage of development. Consequently, there is an evident need for more inclusive studies, such as examining food intake in older adults by comparison with other age groups.

Given the vital role that food intake plays in the health and well-being of older adults [14], and the increasing global burden of diet-related noncommunicable diseases [15], there is a dire need to expand research into how social networks influence dietary behaviors in the senior people. Older adults often face unique nutritional challenges [16] and are more often in situations of increased vulnerability [17] and may benefit from community and social support in maintaining a healthy diet [18]. This is particularly true for the intake of processed foods high in salt, which are linked to hypertension [19], myocardial infarction [20], stroke [21], cancer [22] and other health issues. Moreover, the dietary decisions of older adults are often influenced by relationship-oriented factors such as leisure and cognitive activities, network size, marital status, or changes in social roles [12, 23]. Future research could provide valuable insights into the potential for social network interventions that promote healthier eating habits among older adults [7]. Such interventions could target social norms, support behavior change, and leverage existing relationships to mitigate the risks associated with high salt consumption [24]. Addressing unhealthy food behavior of older adults from a social network perspective may prove beneficial for gaining insights into prevention.

The global population aged 65-year-old plus is projected to increase from 10% (2022) to 16% (2050) [25]. Romania (a European Union (EU) Eastern European country) reports approximately 3.7 million people aged 65 or over (19.2% of the population) and expects to reach 27.7% in 2050 [26]. In 2022, Romania had the fifth highest number in the EU (37.2%) of older adults at risk of poverty or social exclusion [27]. The situation of Romania is not distinct from the rest of the Eastern Europe. The ageing population in Eastern Europe presents a significant demographic shift, characterized by an increasing proportion of older adults due to higher life expectancies and declining birth rates. This trend is leading to several challenges: healthcare systems are strained by the growing need for medical care specific to this age group; the economy faces pressures from a shrinking workforce and the rising demand for pensions and social services; and there is a marked urban-rural divide, with rural areas often having a higher percentage of senior residents.

Older adults living in rural areas of Eastern Europe face similar challenges influencing their health, social life, and overall well-being [28]. Specifically, the seniors have limited access to healthcare, experience high levels of social isolation, rely on inadequate pensions or savings to meet their needs, have lower levels of formal education than their urban peers, and lack the infrastructure essential for comfortable ageing. On top of that, we underline the health disparities. Rural older adults may have higher rates of certain health problems (e.g., unmet dental examination) due to less access to preventive care, lifestyle factors, and the physical demands of rural living [29].

The available literature claims Eastern European countries share not only historical experiences (e.g., the transition from Communism) or trends in population ageing, but also cultural specificities of eating habits. The extant findings show, for instance, that adherence to the traditional Eastern European diet is significantly associated with risks of all-cause and cardiovascular deaths [30].

Despite the acknowledged impact of dietary habits on health outcomes in ageing populations, the intersection of networks and dietary choices in these communities remains underexplored, particularly within rural Eastern Europe’s unique socioeconomic and cultural contexts. In this paper, we investigate the influence of social networks on the consumption of processed food high in salt in community-dwelling older adults and other age-groups, in rural Romania (a representative Eastern European country). We focus our attention on salty food intake given its negative impact on health status and due to the tendency of Eastern European countries to display high levels of sodium intake [31]. In our study, we operationalize the influence of social networks as *assortativity*, i.e., similar people are more likely to be connected or share a tie [32]. We hypothesize that older adults eating processed food high in salt are more likely to be connected to social contacts with similar eating habits. We advance that food intake preferences are clustered in social networks and not randomly distributed. We test this hypothesis on cross-sectional real-world observed social data. We deploy a personal network research design [33] to collect data from a sample of individuals living in a small Romanian rural locality. As our data are not longitudinal, we limit the scope of our study to assortativity detection. Therefore, disentangling the causes of assortativity (social selection, influence, and environmental confounding) is not the object of our current attention. By focusing on older adults from an Eastern European rural area, our research offers valuable insights into the social determinants of dietary behavior, contributing novel data to a previously scant area of gerontological nutrition literature. Our work may enrich the understanding of dietary patterns among rural Eastern European seniors and provide an empirical foundation for designing targeted interventions to mitigate health risks associated with high-salt diets in older populations.

## 2. Methods

### 2.1. Participants

We sampled from a small Romanian rural community (N = 4,557, September 13 – 30, 2023). Lere=ti locality (part of the Romanian county Arge=) is 147 km North-northwest of Bucharest (the capital of Romania). We deployed a personal network research design [33–34]. A number of 83 people (dubbed *egos* in the network science vocabulary; 18 years old plus,) responded to a network questionnaire. They reported information about themselves and their social contacts (maximum 25 *alters* with whom they interact most frequently). We collected both attribute data (socio-demographic and self-declared food intake behavior) and network data (how study participants and their alters interact).

### 2.2. Procedures

Our research is part of the 4P-CAN project (Personalized cancer primary prevention research through citizen participation and digitally enable social innovation; HORIZON-MISS-2022-CANCER-01, project ID 101104432, programme HORIZON). This project addresses the major modifiable risk factors for cancer to improve medical prevention activities and reduce inequalities in Eastern European countries. Our paper contributes to the 4P-CAN research objectives. We collected data in one of the 4P-CAN living labs (Lere=ti). Before initiating our study, we informed the local authorities and community about the 4P-CAN projects’ objectives through press conferences (broadcasted by local media), various meetings with community citizens, authorities and local key actors (video content is available on the project’s web page and social media accounts such as LinkedIn, Facebook and X).

All methods were carried out in accordance with the relevant guidelines and regulations (specifically, those provided by the Romanian Sociologists Society, i.e., the professional association of Romanian sociologists). All the research protocol was approved by a named institutional/licensing committee. Specifically, the Ethics Committee of the Center for Innovation in Medicine (InoMed) reviewed and approved all these study procedures (EC-INOMED Decision No. D001/19-01-2024). Informed consent was obtained from all subjects. The privacy rights of the study participants were observed.

Before each interview, we explicitly informed each participant about the 4P-CAN research objectives. Participants freely signed a consent form and received a paper dossier with informative materials about the project. We granted our participants anonymity and the possibility of opting out at any moment. We informed our respondents that a special web page in Romanian with details about the methodology was specially created for them [35]. We also provided the participants with the contact details (phone numbers and email addresses) of the project director and research team in case they have future queries. We kept total transparency about the project and the research activities to facilitate citizenship engagement in the study.

We gathered the data through face-to-face interviews. On average, an interview had a duration of approximately 80 minutes. We recorded the responses using the Network Canvas software package [36]. Given its flexible architecture, Network Canvas allows for an interactive data collection process wherein participants can see the questions and review their responses in real time.

We did not offer monetary compensation to the study participants. We rewarded them by providing free access to a local educational program focused on health topics and a hotline number in case they need a secondary opinion on medical problems or healthy nutrition.

We employed a link-tracing sampling methodology to recruit participants, i.e., a respondent-driven sampling variant [37, 38]. Due to its expected superior response rates, we used this sampling technique, not a probabilistic non-network random sampling strategy. In a face-validity manner, we considered a network-oriented sampling method more appropriate given the questionnaire size (eventually, our response rate was 56%).

**Fig. 1** illustrates the sampling procedure. We began with six individuals (the seeds). We invited these people to respond to our questionnaire and recommend other interested peers (we did not restrict the possible number of recommendations). We selected the seeds based on ethnographic fieldwork (June – September 2023) that preceded the face-to-face interviews. (During the fieldwork, we explored the community, developed local connections, presented the project to the authorities, took notes about the local realities, etc.). We were interested in having seeds with a different socio-demographic profile (as possible) and located in different geographical points of the community (despite its small size of 140,49 km^2^). Therefore, our seeds were different in sex (four males and two females), age (*Range* = 30, *Min =* 34, *Max* = 64), personal income (three below and three above the average national net salary), employment sector (one retired, three employed in public sector, one employed in private sector, and one self-employed) and education (five with university degree, and one with high-school diploma). Two of the six seeds refused to participate as an interviewee, yet they still made recommendations for participation.

**Fig. 1.**
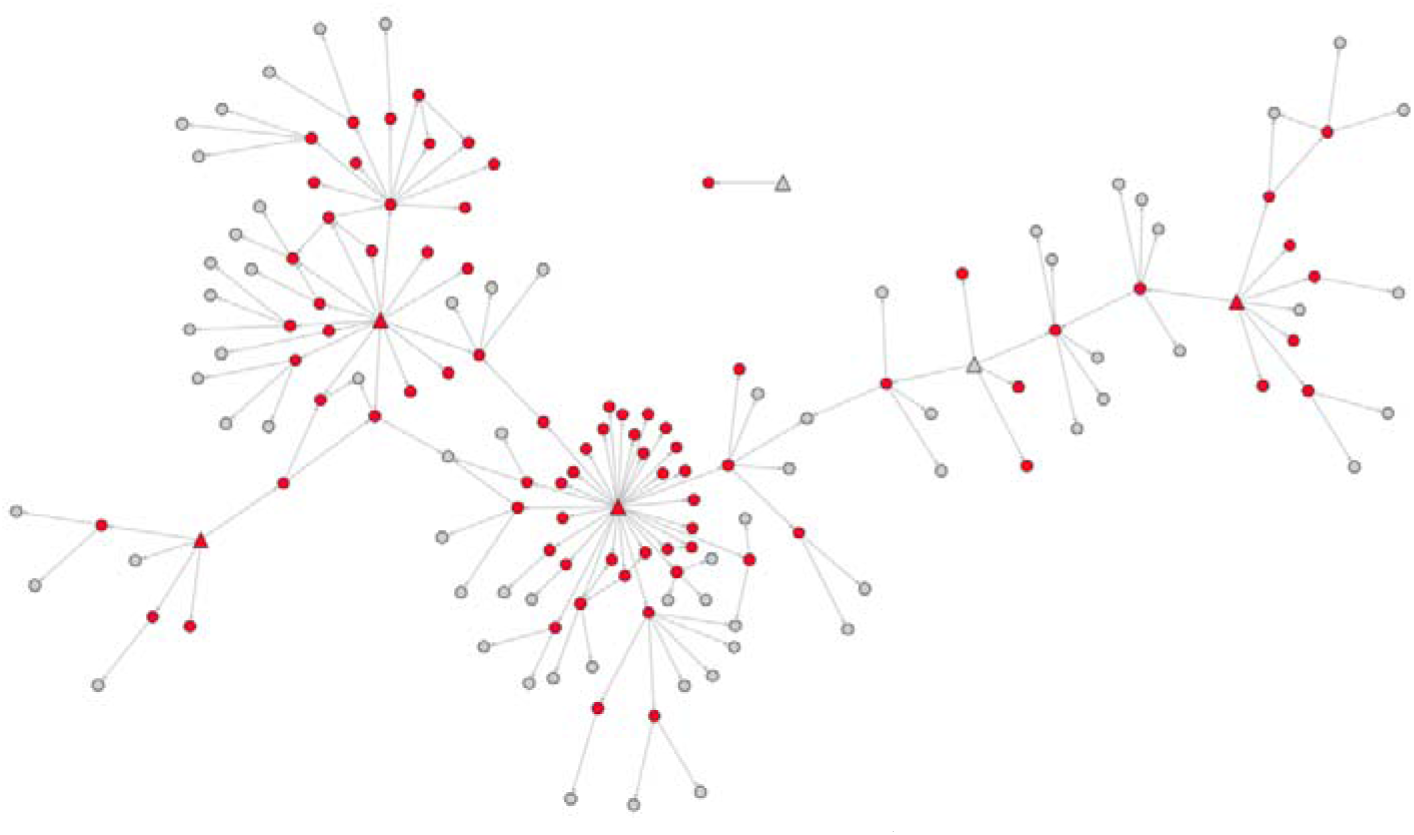
Link-tracing sampling. The triangles mark the seeds (initial people invited by the researchers to participate in the study). The circles indicate people referenced by other study participants. The red indicates participation, while the grey marks non-participation for various reasons (refusals, rescheduling, etc.). The arrows show the referee–referral chains.

After interviewing the seeds, we continued to collect data from the people recommended by each seed and, then, from the persons recommended by the people recommended by the seeds, and so on. Essentially, we followed referee-referral chains as depicted in **Fig. 1**. We stopped the data collection process when our sample (n = 83) had individuals of various socio-demographic profiles: sex (males and females), age (young, adults, and older adults), education (university, high school, elementary), and personal income (below and above the average national net salary).

### 2.3. Data

#### 2.3.1. Network data

We asked each participant (ego) to nominate a maximum of 25 social contacts (alters) [39], i.e., people with whom they interact most frequently (18-year-old plus). *Please nominate 25 people (18 years old plus) you interact with (or meet). You can start with the people you interact with most often. These may be family members, friends, acquaintances, neighbors, work colleagues, etc.* This question allowed us to create a list of ego-alter ties for each participant.

Furthermore, each ego evaluated each pair of alters (alter-alter tie): whether the elicited social contacts know each other personally. And if they do, whether they are *close friends, simple friends,* or *acquaintances*. In this way, we eventually created a personal network for each ego. **Fig. 2** presents two personal networks as an illustration. For each ego-alter tie, we asked our respondents information about the frequency of the meetings (*How often do you typically meet with each of the people you previously mentioned?* Responses: *daily, weekly, every two weeks, once a month, a few times a year, once a year, less than once a year*) and their emotional closeness (*How emotionally close do you feel to the people you previously mentioned?* Responses: *very close, close, not very close, not close at all*). We used this information to estimate the intensity of each ego-alter relationship. First, we dichotomized meeting frequency (Daily meeting = 1, the rest of the responses = 0) and emotional closeness (very close = 1, the rest of the responses = 0). Second, we multiplied the binary variants of meeting frequency and emotional closeness and obtained the intensity of an ego–alter tie.

**Fig. 2.**
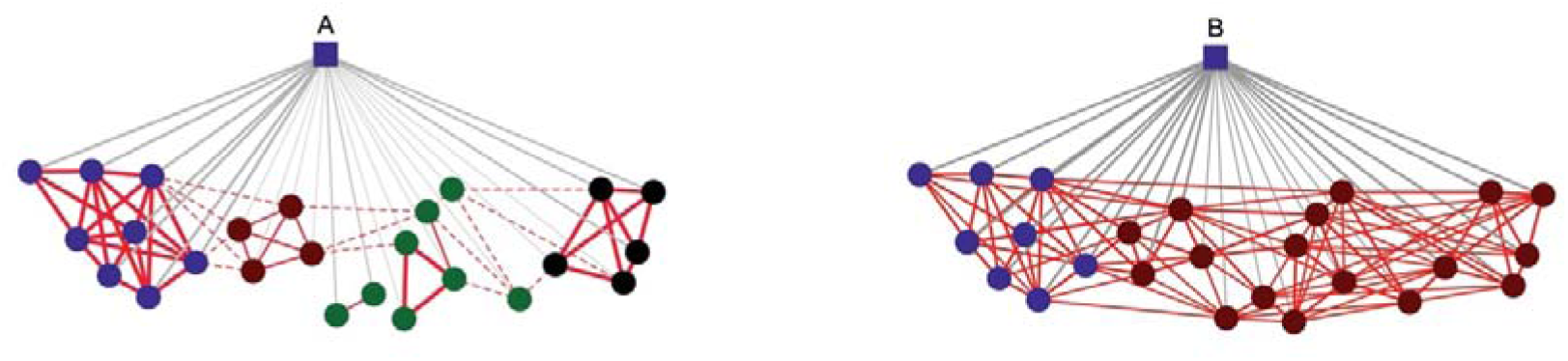
An illustration of two personal networks. The square nodes represent the egos (the study participants), whereas the circles mark the alters (the social contacts elicited by the egos). The colors indicate the alters’ status in relation to the ego: blue (family members), dark red (close friends), green (simple friends), and black (acquaintances). The colors of the ties mark either ego-alter ties (grey) or alter-alter ties (red). The width of grey ties marks the ego-alter meeting frequency (width increases as meeting frequency increases). The width of the red ties marks the type of the alter-alter ties (very thick lines: close friends; simple lines: simple friends; interrupted lines: acquaintances). Images A and B visually illustrate the idea of network density (proportion of materialized alter-alter ties in each personal network). Network A has a lower network density score in comparison to B. Both networks have a size of 25 (this corresponds to the number of alters).

We employed the network information to create two network-level variables for each of the 83 personal networks: the network size (the number of elicited alters) and the network density (the number of observed alter-alter ties divided by the number of theoretically possible alter-alter ties).

#### 2.3.2. Attribute data

Participants provided information about their sex (as assigned at birth: male or female), date of birth, last level of education, occupational status (active or retired), civil status (single or in a relationship), personal income level (above or below the national average net salary) and if they practice any sport (yes/no). The egos gave information about each of their nominated alters: age (in completed years), sex (male or female), education (last level), and whether they were on a diet or had any sport as a hobby (yes/no). By *diet,* we referred to any restrictive eating or a regulated eating plan irrespective of the reasons whereas by *sport* we referred to anything involving physical activity.

We computed the Body Mass Index (BMI) for each ego based on self-declared weight (in Kg) and height (in cm). We divided the weight by the height squared. Then, we classified the resulting scores into “underweight” (less than 18.5), “normal” (18.5 – 24.9), “overweight” (24.9 – 29.9), and “obesity” (higher than 29.9). We also measured our respondents’ perceptions of their alters’ BMI. Namely, we administered pictorial images of females and males [40] to our respondents. We registered the reported corresponding BMI class for each alter (social contact), i.e., ten classes from A to J corresponding to underweight, normal weight, overweight, and obesity classes I, II and III.

We measured the consumption of processed food high in salt (PFHS) by administering a pictorial image to our respondents (See Supplementary Material, Fig 1S; [41]. Each participant reported the frequency of PFHS intake for them and their elicited alters (*How often do you (How often does alter) consume foods similar to the ones depicted in this pictorial image?* Responses: *daily, weekly, every two weeks, once a month, a few times a year, once a year, less than once a year, never*). We dichotomized the responses into a binary variable: whether the ego (alter) eats PFHS food *frequently (daily* or *weekly*). We built the pictorial image using the guidelines available in the WHO STEPS Surveillance Manual [42]. The pictorial image included canned noodle soup, pretzels and chips, burgers, pizza, canned food, and a deli meats platter. We adapted the image to the Romanian cultural specificities by including pickled food and grilled minced meat.

#### 2.3.3. Compositional data

Based on the information reported by the ego, for each personal network, we computed *the proportion of alters* by sex (e.g., proportion of females), age categories (young [18 – 30-year-old], middle [31 – 63 year-old] and older adults [64 year-old and older]), education (e.g., proportion of alters with a university degree), BMI class (e.g., proportion of alters with obesity), diet (proportion of alters on a diet), sport (proportion of alters with a sport as a hobby), PFHS intake (proportion of alters that eat PFHS frequently), closeness (proportion of alters nominated as very emotionally close) and meeting frequency (proportion of alters that ego meets daily). Further, we used closeness and meeting frequency to generate the average intensity of ego-alter ties.

#### 2.3.4. The assortativity variable

We computed a food intake assortativity variable for each alter embedded in a personal (ego) network. Namely, the difference between *the proportion of the alter’s direct contacts who eat PFHS frequently* and *the proportion of alters in the entire ego network that eat PFHS frequently (without the alter of reference)*. (Eating PFHS is a dichotomized variable, where 1 means frequent processed food intake and 0 does not.) The assortativity variable tests whether alters’ PFHS intake associates with the PFHS intake of those to which they are connected (their social contacts). The assortativity variable tells us whether an alter’s direct contacts eat PFHS more or less frequently than all the alters in the ego network. The normalization obtained by subtracting the average PFHS intake of overall alters (minus the alter of reference) in a network is necessary. Without this normalization, the effect of this variable would be confounded by the overall ratio of alters frequently eating PFHS in an ego network. (Variation of PFHS ratios over the different ego networks is controlled for by random ego-level effects). Across the 83 ego networks, we dropped the alters without direct contacts from the analysis. The assortativity variable was undefined in their case. **Fig. 3** visually illustrates the logic behind the assortativity variable. In the displayed personal network, alter *k* has four direct contacts (1, 2, 3, and 4), of which only two eat PFHS frequently (3 and 4). In the entire personal network, there are 24 alters (plus the alter of reference, *k,* that we exclude from the calculations), and six alters eat PFHS frequently (see the red nodes). The assortativity variable would equal 2/4 – 6/24 = 1/4. This result means that the direct contacts of alter *k* have an average PFHS intake behavior above the average PFHS intake behavior in the personal network (they are more likely to eat PFHS frequently than the rest of the alters in the same network). The alter of reference is not included in the computation to avoid circular dependency in the data. Here, we replicate the mathematical formula used to assess assortativity in the case of COVID-19 vaccination [43].

**Fig. 3.**
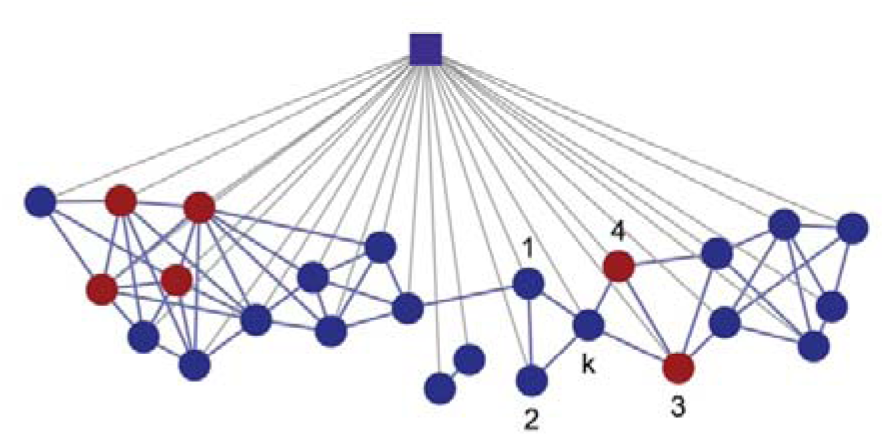
The distribution of processed food high in salt (PFHS) intake behavior in a personal network. The colors indicate eating patterns: red marks people who eat PFHS frequently, whereas blue marks people who eat PFHS less frequently. The geometrical shapes present the ego (the square) and the social contacts (the circles). Alter-alter ties are in blue, and ego-alter ties are in grey. In the personal network, we observe 25 alters and one ego. Alter *K* has four direct contacts (1, 2, 3, and 4).

### 2.4 Statistical analysis

First, we performed a statistical summary to characterize the study participants in the sample (egos) and their social contacts (alters). Second, we fit multi-level, mixed-effects binary logistic regression models, clustering alters by their egos, to predict alters’ PFHS intake behavior (1 = eating PFHS frequently, 0 = eating PFHS less frequently). In these models, we controlled for the attributes of alters (sex, age category, whether they are on a diet, and BMI), of egos (sex, age, education, income, PFHS intake, whether they live single and have a sport as a hobby), and for network properties (network density, network size, and the average intensity of ego-alter ties). We also included the assortativity variable as our key predictor. Third, we fit standard logistic regression models with binomial distribution to predict egos’ PFHS intake behavior (1 = eating PFHS frequently, 0 = eating PFHS less frequently). Again, we controlled for the attributes of egos, for the proportion of alters in the personal networks (females, older adults, alters with obesity and alters on a diet), and for network characteristics (average intensity of ego-alter ties and network density). In the models, we introduced the proportion of alters that eat PFHS frequently as a key variable to detect assortativity. We reported additional details about the regression models (ICC, AIC, BIC, number of observations and pseudo-R^2^) and considered statistically significant effects with *p-values* less than 0.05 from two-sided tests. We prepared detailed diagnostics about the regression models in the Supplementary Material while the data for replication is openly available [41].

In our study, we hypothesized that older adults eating processed food high in salt are more likely to be connected to alters with similar eating habits. We built the statistical models so that we can test the hypothesis.

## 3. Results

### 3.1. Descriptive statistics for the variables of interest

**Table 1** reports the characteristics of the study participants (egos) and their social contacts (alters). The average age of the egos is 54.3 years (standard deviation, *SD*: 15.7). Most of the participants are females (53%), have a personal income below the average national net salary (77%), are not single (87%), and do not have a university degree (18% + 37% = 55%). Also, 54% do not do any sport, and 46% eat PFHS regularly (daily or weekly). Looking at the composition of the 83 personal networks, we note that the average proportion of females is 0.53, of older adults is 0.28, of alters on a diet is 0.13, of alters with obesity is 0.36, and of alters that eat PFHS regularly is 0.43. The egos have relatively dense networks (*M:* 0.7*, SD*: 0.2) and, on average, a network size of 23.7 (*SD*: 3.1).

**Table 1.** Descriptive statistics of the variables of interest. Characteristics of the study participants (egos) and their social contacts.

| The egos (n = 83) |  | The social contacts of the egos (n = 1970) |  |
| --- | --- | --- | --- |
| Variables | Statistics | Variables | Statistics |
| Sex, no. (%) |  | Eating PFHS, no. (%) |  |
| 0 [male] | 39 (47%) | 0 [No] | 941 (48%) |
| 1 [female] | 44 (53%) | 1 [Yes] | 847 (43%) |
| Age, years, mean (SD) NA | 54.3 (15.7) 0 | NA | 182 (9%) |
| Education, no. (%) |  | Sex, no. (%) |  |
| 0 [elementary] | 15 (18%) | 0 [male] | 921 (47%) |
| 1 [high school] | 31 (37%) | 1 [female] | 1049 (53%) |
| 2 [university] | 37 (45%) | Age category, no. (%) |  |
| Single, no. (%) |  | 0 [young: 18 – 29-year-old] | 190 (10%) |
| 0 [No] | 72 (87%) | 1 [middle: 30 – 63-year-old] | 1214 (62%) |
| 1 [Yes] | 11 (13%) | 2 [older adult: 64-year-old +] | 566 (29%) |
| Personal income, no. (%) |  | Diet, no. (%) |  |
| 0 [below national avg.] | 64 (77%) | 0 [No] | 1607 (82%) |
| 1 [above, national avg.] | 17 (20%) | 1 [Yes] | 282 (14%) |
| NA | 2 (3%) | NA | 81 (4%) |
| Doing sport, no. (%) |  | BMI, no. (%) |  |
| 0 [No] | 45 (54%) | 1 [under-weight] | 201 (10%) |
| 1 [Yes] | 38 (46%) | 2 [normal weight] | 284 (14%) |
| Eating PFHS, no. (%) |  | 3 [normal weight] | 405 (21%) |
| 0 [No] | 45 (54%) | 4 [over-weight] | 343 (17%) |
| 1 [Yes] | 38 (46%) | 5 [obesity - I] | 250 (13%) |
| Prop. females, mean (SD), NA | 0.53 (0.19) 0 | 6 [obesity - I] | 181 (9%) |
| Prop. older adults | 0.28 (0.22) 0 | 7 [obesity - II] | 134 (7%) |
| Prop. alters on diet | 0.13 (0.13) 1 | 8 [obesity - II] | 76 (4%) |
| Prop. alters with obesity (BMI) | 0.36 (0.18) 1 | 9 [obesity - III] | 53 (3%) |
| Prop. PFHS intake by alters | 0.43 (0.34) 0 | 10 [obesity - III] | 21 (1%) |
| Avg. intensity of ego-alter ties | 0.12 (0.14) 0 | NA | 22 (1%) |
| Netw. size, mean (SD), NA | 23.7 (3.1) 0 | Ego-alter tie intens., mean (SD), NA | 0.12 (0.33) 0 |
| Netw. density, mean (SD), NA | 0.7 (0.2) 0 | Assortativity, mean (SD), NA | 0.00 (0.1) 192 |

SD = standard deviation. NA = missing data. Netw. = Network. intens. = intensity. Prop. = proportion. PFHS = processed food high in salt. Avg. = average.

Most of the egos’ social contacts are females (53%), overweight and with obesity (17% + 13% + 9% + 7%+ 4% + 3% + 1% = 54%), and not on a diet (82%). A consistent share of the alters eat PFHS regularly (43%) and are older adults (64-year-old and older: 29%). The assortativity variable is positive (*M:* 0.002, *SD*: 0.11). This indicates that, on average, the neighbors of a specific alter have a PFHS intake behavior above the PFHS intake behavior computed for the entire ego network. The average intensity of ego-alter ties across networks is 0.12 (*SD*: 0.33). This value suggests that across the 83 personal networks, we have a small share of alters that egos meet daily and consider very close emotionally.

### 3.2. Predicting the food intake for participants’ social contacts (alters)

**Table 2** shows the results of two multi-level logistic regression models that predict the PFHS intake behavior of the egos’ social contacts (alters). In Model 1, we look at the individuals’ attributes (alters and egos) while controlling for the size of the personal networks (’network size’). We observe that *alter age* and *alter* being on a *diet* are negatively associated with PFHS intake. Our data (1,693 valid observations from 80 personal networks) suggest that younger people are more likely (OR 0.57, *p* 0.00) and individuals on a diet are less likely (OR 0.23, *p* 0.00) to eat PFHS regularly. However, social contacts with higher BMI are perceived as more likely to consume PFHS (OR 1.44, *p* 0.00). Also, the alters embedded in the personal networks of males (OR 0.26, *p* 0.04) and of egos that eat PFHS regularly (OR 4.81, *p* 0.02) are more likely to be perceived as eating PFHS regularly.

**Table 2.** Multi-level logistic regression models. Predicting the intake of processed food high in salt for alters based on the characteristics of egos, alters and network features (assortativity included).

|  | Model 1 |  | Model 2 |  |
| --- | --- | --- | --- | --- |
|  | OR (95% CI) | p | OR (95% CI) | p |
| (Intercept) | 0.79 (0.20 - 3.08) | 0.73 | 0.82 (0.20 - 3.36) | 0.79 |
| *Attributes of alters* |  |  |  |  |
| Sex | 0.86 (0.64 - 1.16) | 0.32 | 0.85 (0.63 - 1.15) | 0.29 |
| Age category | 0.57 (0.48 - 0.68) | 0.00 | 0.58 (0.49 - 0.69) | 0.00 |
| Diet | 0.23 (0.15 - 0.35) | 0.00 | 0.23 (0.15 - 0.35) | 0.00 |
| BMI | 1.44 (1.24 - 1.67) | 0.00 | 1.45 (1.24 - 1.69) | 0.00 |
| *Attributes of egos* |  |  |  |  |
| Sex | 0.26 (0.07 - 0.95) | 0.04 | 0.25 (0.06 - 0.98) | 0.05 |
| Age | 1.05 (0.56 - 2.00) | 0.87 | 1.02 (0.53 - 1.98) | 0.95 |
| Education | 0.55 (0.28 - 1.10) | 0.09 | 0.53 (0.26 - 1.11) | 0.09 |
| Single | 0.72 (0.12 - 4.28) | 0.72 | 0.63 (0.10 - 3.92) | 0.62 |
| Income | 0.79 (0.15 - 4.29) | 0.79 | 0.81 (0.15 - 4.35) | 0.80 |
| Sport | 1.37 (0.38 - 4.94) | 0.63 | 1.38 (0.38 - 4.95) | 0.62 |
| Processed food intake | 4.81 (1.31 - 17.56) | 0.02 | 4.73 (1.27 - 17.54) | 0.02 |
| *Network properties* |  |  |  |  |
| Intensity of ego-alter tie |  |  | 0.95 (0.61 - 1.49) | 0.83 |
| Network density |  |  | 0.92 (0.46 - 1.85) | 0.82 |
| Assortativity effect |  |  | 1.17 (1.03 - 1.32) | 0.01 |
| Network size | 1.02 (0.61 - 1.71) | 0.93 | 1.01 (0.60 - 1.68) | 0.98 |
| ICC | 0.67 |  | 0.66 |  |
| N (observations) | 1693 |  | 1693 |  |
| N (groups: ego-networks) | 80 |  | 80 |  |
| Group-level variation (SD) | 2.56 |  | 2.55 |  |
| AIC | 1546.67 |  | 1546.33 |  |
| BIC | 1622.75 |  | 1638.71 |  |
| Pseudo-$R^2$ (fixed effects) | 0.17 | | 0.17 | |
| Pseudo-$R^2$ (total) | 0.72 | | 0.72 | |

All continuous predictors are mean-centred and scaled by one standard deviation. The outcome variable is in its original units. OR = odds ratio. CI = confidence interval.

In Model 2, we add structural features to evaluate the role of network characteristics in shaping food habits. First of all, the results suggest similar trends in the attributes of the egos and alters. This indicates consistency in the attributes influencing PFHS intake. Alters’ age (OR 0.58, *p* 0.00), diet (OR 0.23, *p* 0.00), and BMI (OR 1.45, *p* 0.00) have (almost) identical contributions to predicting PFHS intake as in Model 1. The same interpretation applies to egos’ sex (OR 0.25, *p* 0.05) and PFHS intake behavior (OR 4.73, *p* 0.02). Second, the assortativity variable positively associates with the dependent variable (OR 1.17, *p* 0.01). If assortativity increases by one standard deviation, then PFHS odds increases by 17%.

The high ICC values in both models (0.67 and 0.66) underscore the substantial impact of the network context, as evidenced by the variability in the PFHS intake behavior across different personal networks (*SD*: 2.56 and 2.55). The fixed pseudo-R^2^ values indicate that fixed effects account for approximately 17% of the variance in the outcome. The total models (the total pseudo-R^2^ values) explain about 72% of the variance, highlighting the substantial contribution of the random effects structure to the model.

### 3.3. Predicting the food intake for participants (egos)

**Table 3** presents two logistic regression models that predict whether study participants eat PFHS regularly. Model 3 examines the effects of the egos’ and alters’ attributes on the binary outcome. The model shows no significant improvement over the null model (χ^2^ (10) 13.12, *p* 0.22) and a modest explanatory power (Cragg-Uhler 0.21). The lack of significant predictors indicates that individual characteristics of egos and alters do not have a distinguishable impact on the dietary behavior of interest. Notably, the propensity to engage in sports activities, suggests a negative association with the frequency of PFHS intake (OR 0.33, *p* 0.04).

**Table 3.** Simple logistic regression models. Predicting the intake of processed food that is high in salt for study participants (egos) based on their attributes, the attributes of their social contacts (alters) and network.

|  | Model 3 |  | Model 4 |  |
| --- | --- | --- | --- | --- |
|  | OR (95% CI) | p | OR (95% CI) | p |
| (Intercept) | 1.60 (0.50 - 5.13) | 0.43 | 2.18 (0.55 - 8.60) | 0.27 |
| *Attributes of egos* |  |  |  |  |
| Sex | 0.39 (0.08 - 1.80) | 0.23 | 0.21 (0.03 - 1.47) | 0.12 |
| Age | 0.92 (0.41 - 2.08) | 0.85 | 1.28 (0.51 - 3.22) | 0.60 |
| Education | 1.14 (0.65 - 2.01) | 0.64 | 1.14 (0.57 - 2.26) | 0.71 |
| Single | 3.22 (0.68 - 15.30) | 0.14 | 2.93 (0.52 - 16.69) | 0.22 |
| Income | 1.31 (0.35 - 4.94) | 0.69 | 1.47 (0.35 - 6.27) | 0.60 |
| Sport | 0.33 (0.11 - 0.95) | 0.04 | 0.24 (0.07 - 0.85) | 0.03 |
| *Ego-network composition (alter attributes)* |  |  |  |  |
| Prop. females | 1.51 (0.71 - 3.25) | 0.29 | 2.28 (0.88 - 5.89) | 0.09 |
| Prop. older adults | 0.63 (0.28 - 1.42) | 0.27 | 0.33 (0.12 - 0.92) | 0.03 |
| Prop. alters with obesity (BMI) | 1.33 (0.78 - 2.27) | 0.29 | 1.53 (0.83 - 2.84) | 0.18 |
| Prop. alters on diet | 1.39 (0.82 - 2.36) | 0.23 | 1.82 (0.97 - 3.42) | 0.06 |
| Prop. PFHS intake |  |  | 2.42 (1.26 - 4.65) | 0.01 |
| *Network properties* |  |  |  |  |
| Intensity of ego-alter ties (avg.) |  |  | 0.65 (0.24 - 1.73) | 0.39 |
| Network density |  |  | 0.43 (0.20 - 0.92) | 0.03 |
| N (observations: egos) | 79 |  | 79 |  |
| AIC | 116.86 |  | 107.31 |  |
| BIC | 142.93 |  | 140.49 |  |
| Pseudo-R<sup>2</sup> (Cragg-Uhler) | 0.21 |  | 0.41 |  |
| | $\chi^2(10) = 13.12$ ($p = 0.22$) | | $\chi^2(13) = 28.67$ ($p = 0.01$) | |

All continuous predictors are mean-centred and scaled by one standard deviation. The outcome variable is in its original units. OR = odds ratio. CI = confidence interval. PFHS intake = proportion of alters that eat PFHS regularly. Prop. = Proportion. avg. = Average.

Model 4 expands upon Model 3 and includes additional variables related to the ego-network structure and the proportion of alters in a network that eats PFHS regularly (another way of measuring assortativity). This model demonstrates a significant improvement over the null model (χ^2^ (13) 28.67, *p* 0.01) and displays greater explanatory power (Cragg-Uhler 0.41). In this model, sports activities are also a significant predictor (OR 0.24, *p* 0.03), reinforcing the potential influence of physical activity on dietary behavior. The presence of seniors within the network (proportion of older adult alters) is also significant (OR 0.33, *p* 0.03), and the network density shows a significant negative association with the outcome (OR 0.43, *p* 0.03). A notable finding is the significant positive effect of the proportion of alters regularly eating PFHS food (Prop. PFHS intake: OR 2.42, *p* 0.01). This finding suggests that egos are substantially more likely to consume PFHS as the number of their social contacts that eat PFHS food regularly increases. The proportion of alters on a diet (Prop. alters on a diet: OR 1.82, *p* 0.06) is not statistically significant.

The comparison of Models M3 and M4 highlights the importance of including network characteristics and alters’ dietary habits to enhance the understanding of PFHS frequency behaviors within ego networks. While individual demographics alone offer limited insight, incorporating network structure and alters’ eating regularity in Model M4 provides a more nuanced understanding of the determinants of dietary behavior.

The consistency in the significance of sports activity across both models suggests a robust relationship between physical activity and dietary behavior, potentially indicating a lifestyle cluster within the ego networks. The findings underscore the need for a multifaceted approach when studying dietary behaviors, accounting for personal attributes and the broader social and network contexts.

## Discussion

Our study explores the influence of social networks on the eating habits of older adults dwelling in rural regions of Eastern Europe, namely Romania. The assortativity effect (Model 2, OR 1.17, *p* 0.01) is statistically significant, highlighting the potential for dietary habits to propagate within social networks, possibly creating clusters of similar eating behaviors. This effect may be more pronounced in older adults, who may rely more heavily on their immediate social circles due to potential limitations in their social interactions. This result is consistent with the concept of homophily in social networks [44–47] or the social contagion of obesity [8]. Also, it underscores the need for interventions that engage with the social environment to promote healthier eating habits [48].

The substantial intra-class correlation coefficients in the multi-level regression models (M1: 0.67 & M2: 0.66) emphasize the impact of network context on PFHS intake behaviour, suggesting that personal networks may significantly shape individuals’ dietary patterns [2]. This insight is of particular relevance when considering the nutritional health of older adults, as it implies that interventions targeting this demographic should extend beyond the individuals to encompass their social environment.

The analysis of egos’ social contacts also reveals a distinct negative association between age and the consumption of PFHS, with older adults (alters) demonstrating a lower propensity for such dietary choices (M1: OR 0.57, *p* 0.00 & M2: OR 0.58, *p* 0.00). This result aligns with the broader literature indicating a generational divide in dietary preferences, as younger individuals have been shown to have a higher inclination towards processed foods [49, 50]. Also, this finding may be influenced by regional economic and availability factors specific to Eastern Europe that merit further investigation.

Additionally, the alters who adhere to a diet are markedly less likely to eat PFHS (M1 & M2: OR 0.23, *p* 0.00), indicating potentially a conscientious shift towards healthier eating habits. This finding resonates with previous work [51] that emphasizes the role of individual agency and family rituals in shaping eating practices. However, our study did not explore the nature of these diets. Future research should uncover whether these are formally prescribed diets, self-directed diets, or culturally prevalent eating patterns and how adherence was achieved.

The positive correlation between BMI and PFHS intake (M1: OR 1.44, *p* 0.00 & M2: OR 1.45, *p* 0.00) raises concerns about the dietary patterns of individuals with a higher body mass index, perhaps caught in a reinforcing cycle of poor nutritional choices. The complex interplay between dietary choices and metabolic health has been extensively documented [52]. Our study is limited to only underscoring the need to investigate the causal pathways linking dietary behaviors and body weight [53].

Turning to the analysis of the egos’ PFHS intake, we suggest that individual characteristics in isolation may not sufficiently explain variations in PFHS consumption. However, introducing network structure measurements improves explanatory power in our logistic regression models. Notably, including the proportion of older adult alters within the network as a significant predictor (M4: OR 0.33, *p* 0.03) highlights the potential role of this demographic in influencing overall network dietary behavior. The positive effect of the proportion of alters that regularly eat PFHS (M4: OR 2.42, *p* 0.01) can suggest a significant likelihood of individuals adapting their eating habits to mirror those of their social contacts who frequently consume PFHS (an assortativity effect). This result is particularly relevant for older adults, who may be more exposed to and influenced by the dietary patterns prevalent within their close-knit social networks.

The robust relationship between physical activity and egos’ dietary behavior, as evidenced by the significance of sports activity across models (M3: OR 0.33, *p* 0.04 & M4: OR 0.24, *p* 0.03), merits attention. This association may reflect a lifestyle pattern within ego networks where healthier physical activity levels correlate with more nutritious dietary choices. Our findings must be contextualized within the broader scientific discourse, particularly considering Eastern Europe’s socio-cultural and economic backdrop [54]. The intersection of physical activity and dietary behavior necessitates a more nuanced future analysis, potentially informed by the available evidence in the field [55], highlighting the synergistic effects of lifestyle factors on health.

There are several limits that the readers should note while interpreting our results. First, our study relies on cross-sectional data, meaning separating social selection and influence in the context of assortativity is impossible. For instance, it cannot be ascertained whether being in a network with higher PFHS intake leads to consuming more processed food (social influence) or if individuals with a preference for PFHS are more likely to form connections with similar dietary habits (social selection). Additionally, cross-sectional designs do not account for periods when processed food intake might be higher or lower (temporal biases). Collecting longitudinal network data in the future can aim to disentangle social selection and influence and improve self-reporting accuracy. Second, the link-tracing sampling of the study participants has inherent selection biases. For example, more health-conscious individuals may be more willing to participate in a study about prevention, which could skew the results. Similar food intake studies focused on East European countries are rare and employ different methodologies [56, 57], which makes result validation almost impossible. Third, an inherent feature of personal network research designs is that egos report data about themselves and their social contacts. While this type of network research design offers valuable insights into understanding food intake, it may be challenging to control for various biases, such as the false consensus effect (the tendency to see one’s own choices replicated in the others) [58] and recall biases (inaccurate reports about PFHS consumption). We tried to manage the false consensus bias by controlling, in our models, the ego-alter tie intensity. Yet, data collected on longitudinal cohorts (balanced panels) can improve the accuracy of self-reported information in the future. Fourth, the generalization of our findings to the entire population is affected by its lack of representativeness.

Our study discusses the impact of assortativity within personal networks on dietary behavior, specifically among older people in rural Eastern Europe who face various vulnerabilities. Consistent with the social integration theory [59], our findings suggest that personal relationships and networks can profoundly impact health behaviors, including diet. It underscores the complexity of dietary choices, shaped by individual characteristics and personal networks (direct alters). A detailed understanding of these social variables is crucial for older adults, whose eating habits are vital for the promotion of healthy ageing and well-being. This knowledge can be utilized to create specific interventions aimed at encouraging better nutrition and cultivating healthier food habits. Community-based interventions can draw inspiration from the social-ecological model of health promotion [60].

## Data Availability

All data produced are available online at https://doi.org/10.5281/zenodo.12703915

## Declarations of interest

none.

## Data availability

The dataset analyzed in the current study is made openly available in the Zenodo data repository as Hâncean, M.-G., Lerner, J., Perc, M., Molina, J.L., Geantă, M., Oană, I. & Pin=oiu-Mihăilă, B.-E. (2024). Replication data for: Processed food intake assortativity in the personal networks of older adults. Zenodo. https://doi.org/10.5281/zenodo.12703915

## Funding

M.-G.H., M.G., I.O., and B.-E.P.-M. were supported by the 4P-CAN project, HORIZON-MISS-2022-CANCER-01, project ID 101104432, programme HORIZON; J.L. was supported by Deutsche Forschungsgemeinschaft (DFG 321869138); M.P. was supported by the Slovenian Research Agency (Javna agencija za raziskovalno dejavnost RS) (Grant Nos. P1-0403). The funding sources had no involvement in study design, in the collection, analysis and interpretation of data, in writing the paper and in the decision to submit the article for publication.

## Acknowledgement

We express our gratitude to Dr. Cosmina Cioroboiu, Dr. Bianca Cuco=, Bogdan-Adrian Vidra=cu, Isabela Tincă, and Dr. Maria-Cristina Ghi=ă for their valuable contributions to this study.

## Author contributions

M.-G.H.: Writing, Visualization, Conceptualization, Methodology, Software, Formal analysis, Investigation, Data curation. J.L.: Writing, Conceptualization, Validation, Methodology, Software, Formal analysis. M.P., J.L.M.: Writing, Conceptualization, Validation. M.G.: Conceptualization, Investigation, Resources, Writing. I.O.: Writing, Investigation, Data curation, Methodology. B.-E.P.-M.: Visualization, Investigation, Data curation, Writing. All authors approved the final version of the paper and accept full responsibility for all aspects of the work described.

## Notes

### Competing Interest Statement

The authors have declared no competing interest.

### Funding Statement

This study was funded by HORIZON-MISS-2022-CANCER-01, project ID 101104432, programme HORIZON; Deutsche Forschungsgemeinschaft (DFG 321869138); the Slovenian Research Agency (Javna agencija za raziskovalno dejavnost RS) (Grant Nos. P1-0403). The funding sources had no involvement in study design, in the collection, analysis and interpretation of data, in writing the paper and in the decision to submit the article for publication.

### Author Declarations

Ethics committee of the Center for Innovation in Medicine (InoMed) gave ethical approval for this work

### Summary of Updates

We have changed the title of the manuscript and the authorship order and composition.

